# Development of a natural language processing application to extract and categorize mentions of violence from mental healthcare records text

**DOI:** 10.64898/2026.03.22.26348435

**Authors:** Lifang Li, Sharon Sondh, Harsharon Kaur Sondh, Robert Stewart, Angus Roberts

## Abstract

**Background:** Experiences of violence are reported frequently by mental health service users, victims of violence are at a greater risk of mental health disorders, and violence may sometimes occur as a consequence of a mental disorder. Electronic health records (EHRs) are an important source of information about healthcare, and its social context. Occurrences of violence are not routinely recorded as structured data in EHRs but are however recorded in the free text narrative.

**Objective:** Our objective was to address this research gap by creating a natural language processing (NLP) application that extracts information related to various forms of violence (physical (non-sexual), sexual, emotional, and financial) from the EHR of a large south London mental health service. Additionally, we aimed to extract features concerning the patient’s role (victimization vs. perpetration), timing (recent vs. historic), domestic context, presence (actual, threat, or unclear), and polarity (affirmed, abstract, or negated) of the violent behaviors.

**Methods:** Two raters independently annotated 6,500 randomly selected segments of clinical notes containing violence-related keywords from a large mental healthcare provider in South London, each containing 400 characters (with approximately 200 characters before and after the keyword) after rigorous training using a pre-defined and approved coding book provided by senior professionals. We utilized 90% of the annotated data for fine-tuning a multi-label BERT model (employing 5-fold cross-validation) with the remaining 10% of data reserved for a blind test.

**Results:** The model performed well on the blind test set for emotional violence (F1= 0.89), financial violence (0.88), physical (non-sexual) violence (0.84), and unspecified violence (0.81), and the patient role (0.89 as perpetrator; 0.84 as victim), polarity (0.89 for affirmed behavior), presence (0.95 for actual violence), and domestic settings (0.88). We were unable to achieve satisfactory results in capturing temporal aspects (0.65 for past violence).

**Conclusions:** We were able to improve substantially on previously developed NLP for ascertaining violence in routine mental health records, providing novel opportunities for both surveillance and research.

## 1. Introduction

The impact of abusive experiences on mental health is profound. Victims of emotional intimate partner violence (IPV) are more susceptible to suicidal ideation, post-traumatic stress, anxiety, depressive symptoms, psychological distress, physical pain, trauma, anger, shame, poor physical health, and somatic symptoms [1]. Specifically, women who have experienced sexual violence during their childhood are more likely to suffer from mental illness in adult life [2]. Victims of domestic violence (DV) may also nurture negative beliefs and attitudes towards others, making it challenging to maintain healthy relationships [3]. Psychological (emotional) abuse has further significant effects on the risk for posttraumatic stress disorder (PTSD) and depression [4]. Additionally, one systematic review found that depression, anxiety, and PTSD were all notable factors associated with both experiencing and perpetrating intimate partner violence [1].

In addition to physical and sexual violence, emotional (psychological) and financial abuse may also be significant components of domestic violence or intimate partner violence [9]. Financial or economic abuse may often go unnoticed, as it occurs within intimate partner relationships [10] [11], and involves tactics such as restricting access to finances, withholding contributions for necessities, limiting access to financial information, and exerting control over household spending [11]. This can have detrimental effects on physical and mental health, as well as on self-esteem and ability to participate in work, social activities, and community engagement [12]. Emotional abuse, a broader experience encountered in daily life, has also proven to be harmful [13], for example as a significant risk factor for partner violence [9, 14], and in the strong correlations of parental emotional abuse with PTSD symptoms in children and adolescents [15], and depressive symptoms in adulthood.

Violence is therefore an important feature of presentations to mental health care. However, there remains a dearth of studies evaluating this from the clinical notes of mental health patients, despite the opportunities offered by these growing digital information resources. In other sectors, NLP has been deployed to open up information resources, notably in the extraction of abuse-related narratives from domestic violence reports provided by the New South Wales Police Force [13]. However, studies to date have often focused on single binary classification models, which may overlook label dependencies and lead to suboptimal results.

To address these research gaps, we applied a multi-label classifier to capture the dependencies and correlations between labels. Also, we sought to develop and improve capability to characterize mentions of violence and abuse from mental healthcare notes using NLP techniques using the Maudsley Clinical Record Interactive Search (CRIS) platform [5]. CRIS was developed in 2008 to enable researcher access to full but de-identified information, within a robust data governance framework, from the electronic health record (EHR) of the South London and Maudsley NHS Foundation Trust, a large provider of mental health care to a catchment population of 1.3m residents in South East London [6,7]. CRIS currently contains data from over 500,000 patients, has supported over 300 published research papers, and has been replicated as a model by several other mental healthcare providers in the UK, expanding the possibilities for multi-site evaluations [8]. Previous work with CRIS resulted in the development of an NLP tool to ascertain violence experiences, sexual abuse, physical abuse, and domestic abuse which was successful but limited in scope [5]. This work reports on the development of a second application, with a greater range of functionality. This redevelopment recognizes that patient role, domestic context, temporality, polarity, and the manifestation of violent behavior are crucial and intersecting features, given that patients may be victims, perpetrators, witnesses, or a combination of these roles [19], and identifying their role is particularly challenging in a domestic context [20]. Our development aimed to provide improved capability for researchers utilizing the CRIS database to explore associations between diverse forms of violence and specific mental health conditions. Furthermore, we anticipated that better recognition of the links between violence and mental health, drawn from research in routine data, might encourage better routine documentation and thus improve information resources as a by-product.

## 2. Methods

### 2.1 Data sources

Our development work comprised two stages. In the first stage, reported in Botelle et al (2022)[5], we collected 3771 instances of potential general violence, sexual violence, and physical violence using the words and word fragments"abus", " assault", " attack", " beat", " chok", " fight", " fought", " hit", " punch", " push", " rape", " slap", " strangl", " struck", " threw", and " violenc". In the second stage, focusing on ascertainment of emotional/financial abuse, text was collected using a search for the following list of word and word fragments “coerciv, emotional abus, emotional manipulat, emotional violen, emotionally abus, gaslight, psychological abus, psychological manipulat, psychological violen, psychologically abus, economic abus, financial abus, financially abus, battered, maltreated”. For each of these strings, we randomly extracted instances up to a maximum number of 300. This resulted in 3500 instances. For these two data collection stages, we excluded resulting in 6,685 instances.

### 2.2 Data labelling

The 6685 instances collected in the previous step are potential mentions of violence. Some of these instances, however, will not be true mentions of violence, having resulted from ambiguous use of the matching strings and keywords. In order to create a dataset for model training and testing, all instances were therefore manually labeled, i.e. annotated, as to the presence or absence of different types and features of violence.

#### 2.2.1 Annotation guidelines

Prior to annotation, we created annotation guidelines aligned with our research objectives, and which defined:

● Violence Types: We provided clear definitions for different types of violence, including emotional, financial, and others. Additionally, we furnished illustrative examples for each violence category.
● Patient Roles and Context: We elucidated the roles that patients could assume—victim, perpetrator, witness, or a combination thereof. Furthermore, we addressed the polarity (affirmed, negated, or abstract), presence (actual, threatened, or unclear), temporality (present, past, or uncertain), and setting (domestic, non-domestic, or ambiguous) associated with violence mentions.

These guidelines served as the foundation for our annotation process, providing consistency and rigor in our data collection. Specific definitions of each violence type and feature are summarized in **Table 1**.

**Table 1.** Definitions of violence types and features.

|  | Values | Definition and explanations | Examples |
| --- | --- | --- | --- |
| Violence type | Emotional | We used the UN definition of psychological violence to define emotional abuse [23]: “Psychological violence encompasses a spectrum of behaviors, spanning acts of emotional abuse and controlling.”<br>Explanation of emotional abuse: it includes insulting, belittling or humiliating, deliberately scaring or intimidating, threatening to hurt the victim or others he/she cares about.<br>Controlling behavior may include the following: isolating the victim; monitoring the victim's whereabouts and social interactions; ignoring the victim; getting angry if the victim speaks with others; making unwarranted accusations of infidelity; controlling the victim’s access to health care, education or labor market. | For example, “We are concerned that relatives are trying to coerce zzzz to do this.” and “Perceived gaslighting” were all affirmed emotional abuse. |
|  | Financial | We used the UN definition of economic violence to describe financial abuse [23]:<br>“Economic violence is said to occur when an individual denies his intimate partner access to financial resources, typically as a form of abuse or control or to isolate her or to impose other adverse consequences to her well-being.”<br>Explanation of financial abuse( economic violence):<br>Economic violence includes the following:<br>restricting the victim's access to financial resources;<br>restricting the victim's access to property and durable goods; intentionally failing to meet economic responsibilities, such as alimony or financial support for the family, thereby subjecting the victim to poverty and hardship; limiting the victim's access to the labor market and educational opportunities;<br>excluding the victim from participation in decision-making processes relevant to economic matters. | “ZZZZZ has suffered from financial abuse.” indicate that the zzzzz is the victim of financial abuse and the financial abuse is affirmed. |
|  | Physical (non-sexual) | We used the UN Home office (2020)’s definition of physical violence [30]. Physical violence comprised violence or abuse that used physical force, or resulted in or had a high likelihood of resulting in physical injury to the victim, but which was not sexual. Thresholds for considering violence physical are partially adapted from UK law, for instance “offenses against the person” (1861 Act) describes crimes which are committed by direct physical harm or force and include spitting as a form of assault and poisoning as a non-fatal, non-sexual offense. | “Tried to hit” indicates the occurrence of physical (non-sexual) violence. |
|  | Sexual | We used the UN Home office (2020)’s definition of sexual violence [30]. Sexual violence included any unwanted sexual act, unwanted sexual comments or advances, unwanted attempts to obtain a sexual act, and directed acts against a person’s sexuality using coercion and acts to traffic. This includes (but is not limited to) rape, sexual harassment, sexual assault, | “ZZZZZ was raped when zzzz was zzzz years old.” This example indicated that the zzzz was the victim of the sexual |
|  |  | forced marriage, FGM, and reproductive coercion and control. | abuse (affirmed sexual abuse). |
|  | Unspecified | If violence occurred, but the type of violence was not specified, or did not fit in one of the types defined above, then the type was given as Unspecified. | “ZZZZ was abused as a zzzz by his zzzz.” General description of being “abused”, not sure which specific type it was. |
|  | Irrelevant | The instance was not about a violence experience pertaining to the patient. | “ZZZZ showed sexually inappropriate behaviour”, this was not about any violence behavior. |
| Patient role (multiple roles allowable) | Perpetrator | Where the patient conducted the violence towards others | “ZZZZ made threats to kill and attack zzzz”. This example indicates that the zzzz is the perpetrator of physical abuse. |
|  | Victim | Where the patient was in receipt of the violent behavior | “ZZZ was abused as a child by his zzzz.” This example indicates that the zzz was the victim of the unspecified violence. |
|  | Witness | Where the patient was not a perpetrator or a victim, but saw/heard the violent behavior | “The zzzzz saw zzz being punched by zzz.” This example indicates that the zzz was the witness of the physical abuse behavior. |
|  | Unclear | Where the patient’s role was unclear |  |
| Polarity | Affirmed | The highlighted violence was described as occurring, | “Ongoing emotional and **financial** abuse.” This example suggested affirmed emotional abuse and financial abuse. |
|  | Abstract | The highlighted violence was being conjectured. Risk and hypothetical violence should be labeled with a polarity of abstract. | “ZZZZ am concerned that there could be financial abuse.” This is one example that mentions hypothetical violence. |
|  | Negated | The highlighted violence was being described as not happening | “No **violence** or aggression noted”. |
| Presence | Actual | A violent act was being described | “ZZZZ hit the zzzz” indicates the actual physical violence. |
|  | Threat | The violence constituted a threat | “ZZZZ made a gesture as if to attack zzzz” this indicates the threatening behavior rather than an actual one. |
|  | Unclear | It was not clear whether the violence was actual or threatened |  |
| Temporality | Past | We defined “Past” violence as having occurred more than a year ago. These values were only used if there was clear evidence of when the violence occurred. Importantly, that evidence had to be contained in the text viewable and not from some other part of the record. |  |
|  | Not-past | If the evidence was not clear, then the value of “Not-past” was used. |  |
| Setting | Domestic | Domestic violence comprised violence between family members, intimate partners, ex-intimate partners and household members.<br>The definition of domestic violence was based on the UK government domestic abuse statutory guidance framework 2020 (updated version on 2023) [31], but adjusted to be applicable to all ages and included people who were household members through choice (but not those in institutional, residential, detention or in-patient settings). | “Abused as a zzzz by their zzzz” indicates the domestic setting of the unspecified abuse. |
|  | Not domestic | The violence did not happen in domestic settings and was not referred to as involving family members, intimate partners, ex-intimate partners or household members. | “Abuse zzzz suffered whilst in zzzz” this indicates that the abusive behavior happened between the zzzz and the zzz rather than in a domestic setting. |
|  | Unclear | It was not possible to ascertain the setting of the violent behavior. | “ZZZZ was abused” which has not been mentioned about the setting. |

We list some examples and the corresponding violence type and values in **Table 2**.

**Table 2.** Examples of text fragments, with matched strings, alongside the assigned annotations. (Note that these do not reproduce original clinical text verbatim.)

| Example of text fragment | Violence Type | Patient role | Polarity | Presence | Temporality | Setting |
| --- | --- | --- | --- | --- | --- | --- |
| The zzz said that zzzz partner plays mind tricks and *emotionally manipulates* zzzz. | Emotional | Victim | Affirmed | Actual | Not-past | Domestic |
| ZZZZ said that zzz child was exposed to *psychological abuse* when zzzz left home. | Irrelevant | / | / | / | / | / |
| No evidence of *emotional abuse* | Emotional | / | Negated | / | / | / |
| ZZZZ was *financially abused* by zzz partner when zzzz was zzz. | Financial | Victim | Affirmed | Actual | Past | Domestic |
| ZZZZ said that zzzz had witnessed zzz mother being *psychologically abused* when zzzz was a zzzz. | Emotional | Witness | Affirmed | Actual | Past | Domestic |
| Was *coerced* into taking part against zzzz will. | Emotional | Victim | Affirmed | Actual | Not-past | Unclear |
| ZZZZZ was sexually *abused* several years ago in a past relationship. | Sexual | Victim | Affirmed | Actual | Past | Domestic |
| ZZZZZ says zzz was *beaten* by zzzz boyfriend. | Physical | Victim | Affirmed | Actual | Not-past | Domestic |

#### 2.2.2 Annotation

We used <u>Label Studio</u> to annotate the instances. Out of all the instances, 185 were randomly chosen for a pilot study, aimed at familiarizing annotators with the annotation guidelines. We randomly selected 700 instances for double-annotation from the remaining 6316 instances. Of these 700 instances, 643 underwent successful double-annotation, and the remaining 57 instances were reserved for wider discussion to resolve disagreements. In this round of double-annotation, 372 instances exhibited at least one discrepancy across six annotation categories. To address this, we conducted an additional round of double-checking, where two annotators collaborated to reach a final decision on these instances. All 700 instances from this double annotation step with resolved consistency were later used in developing the NLP model.

For the remaining 5,800 instances, we conducted single annotations in the majority, with 20% double-annotated overlap agreement checks (n=1,008 instances). For 691 instances with at least one discrepancy across six annotation categories, we conducted a double-check as before. Subsequently, all 5,800 instances with resolved consistency from this step were later used to develop the NLP model. In summary, a total of 6,500 instances were annotated and used in NLP model development.

#### 2.2.3 NLP model development

We utilized a pre-trained BERT model as the foundation for NLP model development. This pre-trained model was further fine-tuned using 90% of the annotated datasets, with the remaining 10% of the instances set aside for the blind test. The training data and the blind test data were balanced according to the ethnicity, gender, and age groups of the patients from which they were extracted.

Dataset balancing was based on previous studies finding significant associations of gender and ethnicity with emotional violence and IPV [28], as well as DV experiences and the need for social support [29]. As a result, the training dataset consisted of 5849 instances and the blind test dataset consisted of 651 instances.

The BERT for Multi-label Sequence Classification model was used. It was trained for violence type, patient role, polarity, presence, temporality, and setting. The annotated data were divided into two sets before training the model. During training, we applied 5-fold cross-validation to find model performance (Bert tokenizer, max_lenth=256, and 4 epochs for each fold; the 4th epoch results in each fold achieved the best performance when using AUROC as the evaluation matrix). To make full use of the annotated training dataset, we trained a full model on all the training data for practical use, and evaluated this with the blind test set. For detailed data processing procedures, see **Figure 1**.

**Figure 1.**
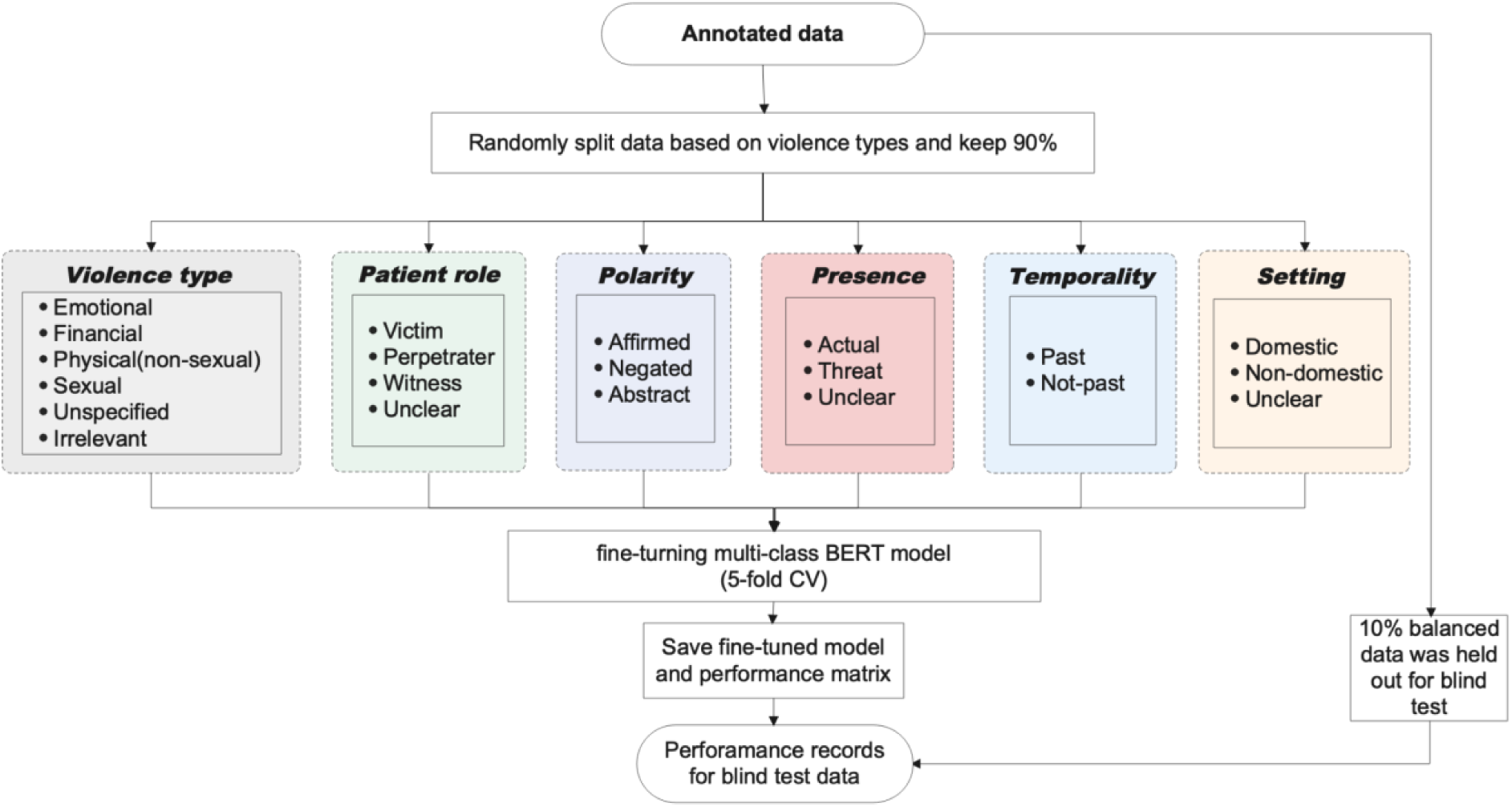
Data processing.

## 3. Results

### 3.1 Annotation agreement

**Table 3** summarizes agreement for double annotation instances. Inter-annotator agreement (IAA) and Cohen’s kappa value demonstrate robust agreement between the two annotators. Specifically, Cohen’s kappa values ranged from 0.78 to 0.84, indicating substantial concordance.

**Table 3.** Agreement levels for double annotations.

| Attribute | Initial double-annotation |  | Random 20% double-annotation |  |
| --- | --- | --- | --- | --- |
|  | IAA | Cohen's Kappa | IAA | Cohen's Kappa |
| Violence type | 0.89 | 0.75 | 0.92 | 0.88 |
| Polarity | 0.84 | 0.76 | 0.89 | 0.83 |
| Presence | 0.85 | 0.76 | 0.88 | 0.82 |
| Patient role | 0.83 | 0.71 | 0.84 | 0.78 |
| Setting | 0.82 | 0.75 | 0.84 | 0.77 |
| Temporality | 0.83 | 0.70 | 0.84 | 0.75 |

### 3.2 Data summary

Of all 6500 instances, 5849 were applied for training and validating and 651 (10%) remained for the blind test. **Table 4** summarizes the datasets and **Table 5** summarizes label frequency in the training and test sets.

**Table 4.** Summary of datasets.

|  | Value | Number in training set | Number in lind test set |
| --- | --- | --- | --- |
| Gender | Male | 2348 | 331 |
|  | Female | 2415 | 319 |
|  | Other | 12 | 1 |
| Ethnicity | White | 2211 | 306 |
|  | Other/NK | 1500 | 202 |
|  | Black | 974 | 131 |
|  | Asian | 90 | 12 |
| Age group | Tertile 1 | 1570 | 225 |
|  | Tertile 2 | 1594 | 216 |
|  | Tertile 3 | 1611 | 210 |

**Table 5.**
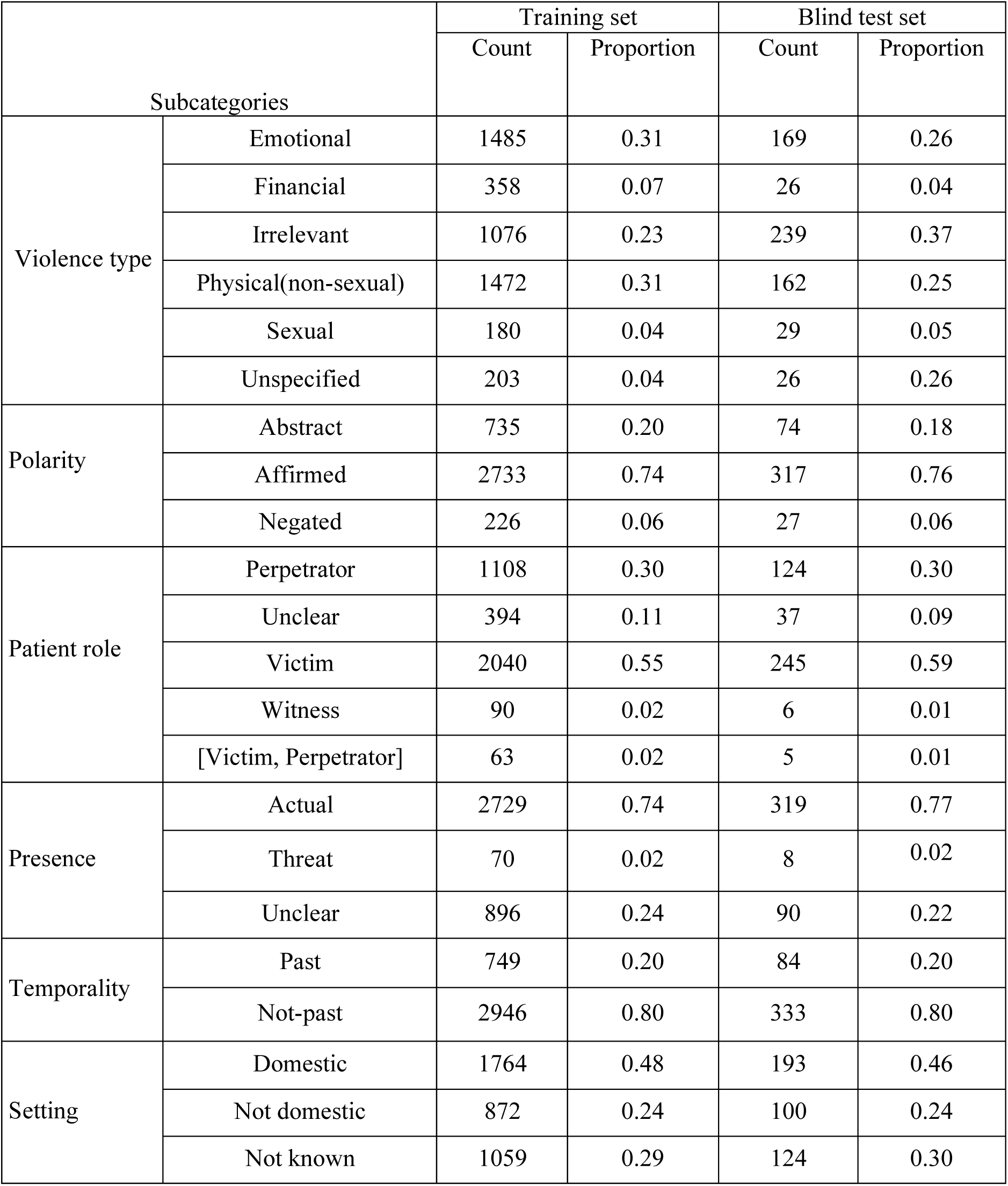
Summary for violence type polarity, patient role, presence, temporality, and setting in training data and blind test data.

### 3.3 Model performance on validation and blind test datasets

**Table 6** summarizes the results of 5-fold cross-validation with 5849 instances as training and validation data. **Table 7** summarizes the model performance on the blind test dataset.

**Table 6:** 5-fold cross-validation results.

|  |  | 5-fold classification (performance on validation dataset) |  |  |
| --- | --- | --- | --- | --- |
|  |  | Precision<br>Mean [95%CI] | Recall<br>Mean [95%CI] | F1<br>Mean [95%CI] |
| Violence type | Emotional | 0.87[0.83,0.91] | 0.97[0.96,0.98] | 0.91[0.89,0.93] |
|  | Financial | 0.96[0.93,0.99] | 0.95[0.94,0.96] | 0.96[0.94,0.98] |
|  | Irrelevant | 0.89[0.88,0.90] | 0.77[0.74,0.80] | 0.82[0.8,0.84] |
|  | Physical(non-sexual) | 0.86[0.84,0.88] | 0.92[0.90,0.94] | 0.89[0.88,0.90] |
|  | Sexual | 0.85[0.80,0.90] | 0.48[0.38,0.58] | 0.61[0.53,0.69] |
|  | Unspecified | 0.94[0.91,0.97] | 0.7[0.64,0.76] | 0.80[0.75,0.85] |
| Patient role | Perpetrator | 0.81[0.79,0.83] | 0.88[0.86,0.90] | 0.84[0.82,0.86] |
|  | Unclear | 0.00[0.00,0.00] | 0.00[0.00,0.00] | 0.00[0.00,0.00] |
|  | Victim | 0.77[0.76,0.78] | 0.9[0.89,0.91] | 0.83[0.82,0.84] |
|  | Witness | 0.00[0.00,0.00] | 0.00[0.00,0.00] | 0.00[0.00,0.00] |
|  | ['Victim', 'Perpetrator'] | 0.00[0.00,0.00] | 0.00[0.00,0.00] | 0.00[0.00,0.00] |
| Polarity | Abstract | 0.81[0.8,0.82] | 0.83[0.83,0.83] | 0.82[0.82,0.82] |
|  | Affirmed | 0.89[0.88,0.90] | 0.91[0.9,0.92] | 0.9[0.89,0.91] |
|  | Negated | 0.93[0.86,1.00] | 0.45[0.32,0.58] | 0.6[0.48,0.72] |
| Presence | Actual | 0.94[0.94,0.94] | 0.93[0.92,0.94] | 0.94[0.94,0.94] |
|  | Unclear | 0.72[0.69,0.75] | 0.75[0.74,0.76] | 0.73[0.71,0.75] |
| Temporality | Past | 0.74[0.71,0.77] | 0.57[0.50,0.64] | 0.62[0.55,0.69] |
|  | Not-past | 0.92[0.91,0.93] | 0.96[0.95,0.97] | 0.94[0.94,0.94] |
| Setting | Domestic | 0.84[0.82,0.86] | 0.9[0.88,0.92] | 0.87[0.86,0.88] |
|  | Not domestic | 0.68[0.63,0.73] | 0.6[0.56,0.64] | 0.63[0.6,0.66] |
|  | Unclear | 0.63[0.61,0.65] | 0.58[0.51,0.65] | 0.6[0.56,0.64] |

**Table 7.** Performance of the BERT base model on the blind test dataset.

|  |  | Blind test (651 instances) |  |  | Count of predicted types |
| --- | --- | --- | --- | --- | --- |
|  |  | Precision | Recall | F1 |  |
| Violence type | Emotional | 0.83 | 0.95 | 0.89 | 169 |
|  | Financial | 0.81 | 0.96 | 0.88 | 26 |
|  | Irrelevant | 0.94 | 0.78 | 0.85 | 239 |
|  | Physical(non-sexual) | 0.78 | 0.91 | 0.84 | 162 |
|  | Sexual | 0.89 | 0.55 | 0.68 | 29 |
|  | Unspecified | 0.90 | 0.73 | 0.81 | 26 |
| Patient role | Perpetrator | 0.87 | 0.91 | 0.89 | 358 |
|  | Unclear | 0.50 | 0.03 | 0.05 | 37 |
|  | Victim | 0.80 | 0.89 | 0.84 | 246 |
|  | Witness | 0.00 | 0.00 | 0.00 | 0 |
|  | ['Victim', 'Perpetrator'] | 0.00 | 0.00 | 0.00 | 0 |
| Polarity | Abstract | 0.88 | 0.86 | 0.87 | 306 |
|  | Affirmed | 0.87 | 0.91 | 0.89 | 318 |
|  | Negated | 0.94 | 0.63 | 0.76 | 27 |
| Presence | Actual | 0.96 | 0.93 | 0.95 | 561 |
|  | Unclear | 0.64 | 0.76 | 0.69 | 90 |
| Temporality | Past | 0.65 | 0.64 | 0.65 | 84 |
|  | Not-past | 0.95 | 0.95 | 0.95 | 567 |
| Setting | Domestic | 0.89 | 0.88 | 0.88 | 427 |
|  | Not domestic | 0.67 | 0.69 | 0.68 | 101 |
|  | Unclear | 0.64 | 0.64 | 0.64 | 123 |

**Table 6** indicates strong model performance in identifying emotional violence, financial violence, physical violence (non-sexual), irrelevant instances, and unspecified violence. However, regarding sexual violence, while precision was high (mean=0.85), recall was not as satisfactory. The model effectively distinguished between perpetrator and victim roles among patients, as well as the polarity of affirmed and abstract violence. Additionally, it accurately detected the presence of actual violence, temporal aspects labeled as ‘Not-past’, and instances of domestic violence. **Table 7** findings were similar, showing that the classification performance for violence type is good. Specifically, on the blind test data the model could correctly identify emotional violence with F1=0.89, financial violence with F1=0.88, physical (non-sexual) violence with F1=0.84, sexual violence with F1=0.67, and unspecified violence with F1=0.81.

Considering the patient role, perpetrator and victim were ascertained with good performance (F1 0.89 and 0.84 respectively). However, performance was unsatisfactory for witnessed violence, and the co-occurrence of various patient roles due to insufficient representation. For polarity, the model could predict abstract violence and affirmed violent behavior well with F1 scores larger than 0.89. For presence, this model performed well in identifying the actual violent experiences of the patients (F1=0.95). Temporality prediction was challenging for the model, as indicated by low precision, recall, and F1 score of the blind test set. The model showed good performance for predicting the domestic setting (F1=0.88).

## 4. Discussion

### 4.1 Principal findings

Following an initial development of NLP to ascertain violence experiences from mental health records [5], this study developed new models with greater functionality. Our objective was to extend detectable violence types, from physical and sexual to encompass emotional and financial abuse, and to develop type-specific characterisations of the patient role (victim, perpetrator, witness), polarity (affirmed, negated, abstract), presence (actual, threat, unclear), temporality (past, not-past, unclear), and setting (domestic, not domestic, unclear). Using 6500 annotated instances we were able to fine-tune a multi-label BERT model with promising results in nearly all respects where there were sufficient annotations available, aside from temporality.

This study describes what we believe to be an important innovation in applying NLP to extract information about violence. Although previous NLP development has been described that extracts information on physical assault, threat, sexual assault, emotional/verbal abuse, stalking, financial abuse, social abuse, and property damage, this was developed for text from police reports and not from health records [13]. Furthermore, NLP previously developed for extracting violence-related information from the Maudsley CRIS mental healthcare data resource was focused on mentions of violence, domestic violence, sexual violence, physical violence, perpetration and victimisation as discrete entities without any functionality to combine these features - for example, to ascertain receipt of domestic sexual violence; in addition, financial and emotional violence were not previously captured [5]. The developments presented here address this research gap and applying the results of this model should have a high potential to transform the ascertainment of violence as an exposure and outcome in mental healthcare, allowing novel research in this sector.

Considering methodology, existing studies have tended to focus on single binary classification models. Training binary classifiers independently may not account for label dependencies, potentially leading to suboptimal results. In contrast, the multi-label classifier we have trained can capture dependencies and correlations between labels. Integrating this information is important for characterizing types of violence [16,26] - most importantly in distinguishing between receipt (victimization) and perpetration of violence, as well as the setting, and other features, all of which are important in the interpretation and application of NLP-derived data in research. For example, it has been found that female emotional abuse victims experiencing abuse more than once a week have an increased risk of depression [27]. Applying the NLP developed here across large data resources will enable further investigation of these exposures as predictors of mental disorder course and outcome.

### 4.2 Limitations

Although successful in many respects, our attempts to create an NLP model that could characterize references to temporality, and capture witnessed violence faced significant challenges. In part, this endeavor was hampered by insufficient sample size, particularly for witnessed violence, preventing the successful development of the intended model. Also, because health records text is inevitably recorded by clinicians in retrospect, the near-universal use of the past tense limited ascertainment of temporality, compounded by text on timing which was either absent or too highly varied for accurate capture; inter-rater agreement was also the lowest for temporality as a construct in both evaluations which may reflect ambiguity in its description.

In addition, we focused on four specific types of violence namely physical (non-sexual) violence, sexual violence, emotional violence, and financial violence, with a further unspecified violence category to capture other types. This characterisation might be improved by adding more types of violence salient to mental health, as well as potentially by more granular categorisation within each type and better distinguishing between threats and actions of violence. In annotations, we sought to assign single violence types for each instance without considering the co-occurrence of various types of violence in one description. In this respect, emotional abuse may co-occur with threats and/or acts of physical and/or sexual violence from intimate partners, constituting acts of violence in their own right. Research indicates that the deployment of various forms of psychological violence is linked to a heightened likelihood of physical and sexual violence against female partners, multifaceted forms of abuse that can significantly impact women, irrespective of whether other forms of violence are present [23]. Future studies could try to overcome this by allowing multiple labels; however, of the 6500 instances annotated in our development, less than 10 described co-occurring types.

As a final consideration particularly applicable to mental healthcare text, in our annotation process, no attempt was made to infer the truth or not of recorded violence instances. Thus, delusional or hallucinated experiences of violent acts or threats were annotated in the same way as non-delusional experiences, as they tended to be described as such in records with only more distant, qualifying text indicating the suspected origin as a false belief or perception. This will be an important consideration when inferences are made from cross-diagnostic prevalence studies and further development would be required for this characterisation to be deployed.

## 5. Conclusion

In this study, we sought to improve capabilities in NLP for structured data extraction of received and perpetrated violence in mental health electronic health records. We were successful in developing a suite of NLP applications using a BERT model to extract information on various types of violence (physical, sexual, emotional, financial) and to characterize the polarity, presence, patient role, and setting. Ascertaining the temporality of the described violence in the same model was more challenging and needs further development. The model performed well in other respects and holds the potential to advance both research and clinical practice.

## Data Availability

The data analysed in this study is subject to the following licenses/restrictions: All the relevant aggregate data are found within the article. The data used in this work have been obtained from CRIS. It provides authorised researchers regulated access to anonymised information extracted from SLaM's electronic clinical records system. Individual-level data are restricted in accordance with the strict patient-led governance. Data are available for researchers who meet the criteria for access to this restricted data: (i) SLaM employees or (ii) those having an honorary contract or letter of access from the trust. Requests to access these datasets should be directed to CRIS administrator

## Acknowledgements

RS is part-funded by: i) the NIHR Maudsley Biomedical Research Centre at the South London and Maudsley NHS Foundation Trust and King’s College London; ii) the National Institute for Health Research (NIHR) Applied Research Collaboration South London (NIHR ARC South London) at King’s College Hospital NHS Foundation Trust; iii) UKRI – Medical Research Council through the DATAMIND HDR UK Mental Health Data Hub (MRC reference: MR/W014386); iv) the UK Prevention Research Partnership (Violence, Health and Society; MR-VO49879/1), an initiative funded by UK Research and Innovation Councils, the Department of Health and Social Care (England) and the UK devolved administrations, and leading health research charities; v) the NIHR HealthTech Research Centre in Brain Health at King’s College London and South London and Maudsley NHS Foundation Trust.

AR is funded by i) Health Data Research UK, an initiative funded by UK Research and Innovation, Department of Health and Social Care (England); ii) devolved administrations, and leading medical research charities and iii) the UK Prevention Research Partnership (Violence, Health and Society; MR-VO49879/1), an initiative funded by UK Research and Innovation Councils, the Department of Health and Social Care (England) and the UK devolved administrations, and leading health research charities.

HKS is a fully funded PhD student at the National Institute for Health Research (NIHR) Maudsley Biomedical Research Centre.

SS is a fully funded PhD student at the NIHR HealthTech Research Centre in Brain Health.

This research was supported by the UK Prevention Research Partnership (Violence, Health and Society; MR-VO49879/1), which is funded by the British Heart Foundation, Chief Scientist Office of the Scottish Government Health and Social Care Directorates, Engineering and Physical Sciences Research Council, Economic and Social Research Council, Health and Social Care Research and Development Division (Welsh Government), Medical Research Council, National Institute for Health and Care Research, Natural Environment Research Council, Public Health Agency (Northern Ireland), The Health Foundation, and Wellcome. The views expressed in this Article are those of the authors and not necessarily those of the UK Prevention Research Partnership or any other funder.

## References

1. Spencer, C., Mallory, A. B., Cafferky, B. M., Kimmes, J. G., Beck, A. R., & Stith, S. M. (2019). Mental health factors and intimate partner violence perpetration and victimization: A meta-analysis. Psychology of Violence, 9(1), 1–17. 10.1037/vio0000156

2. Mullen PE, Martin JL, Anderson JC, Romans SE, Herbison GP. Childhood sexual abuse and mental health in adult life. Br J Psychiatry 1993;163(DEC.):721–732. PMID:8306113

3. Kendall-Tackett K. The health effects of childhood abuse: Four pathways by which abuse can influence health. Child Abus Negl 2002;26(6–7):715–729. PMID:12201164

4. Nathanson AM, Shorey RC, Tirone V, Rhatigan DL. The Prevalence of Mental Health Disorders in a Community Sample of Female Victims of Intimate Partner Violence. Partner Abuse 2012;3(1):59–75. [doi: 10.1891/1946-6560.3.1.59]

5. Botelle R, Bhavsar V, Kadra-Scalzo G, Mascio A, Williams M V., Roberts A, Velupillai S, Stewart R. Can natural language processing models extract and classify instances of interpersonal violence in mental healthcare electronic records: An applied evaluative study. BMJ Open 2022;12(2):1–10. PMID:35172999

6. Vaci N, Liu Q, Kormilitzin A, De Crescenzo F, Kurtulmus A, Harvey J, O’Dell B, Innocent S, Tomlinson A, Cipriani A, Nevado-Holgado A. Natural language processing for structuring clinical text data on depression using UK-CRIS. Evid Based Ment Health 2020;23(1):21–26. PMID:32046989

7. Chilman N, Song X, Roberts A, Tolani E, Stewart R, Chui Z, Birnie K, Harber-Aschan L, Gazard B, Chandran D, Sanyal J, Hatch S, Kolliakou A, Das-Munshi J. Text mining occupations from the mental health electronic health record: A natural language processing approach using records from the Clinical Record Interactive Search (CRIS) platform in south London, UK. BMJ Open 2021;11(3):1–11. PMID:33766838

8. Stewart R, Velupillai S. Applied natural language processing in mental health big data. Neuropsychopharmacology [Internet] Springer US; 2021;46(1):252–253. PMID:32895453

9. Fulu E, Jewkes R, Roselli T, Garcia-Moreno C. Prevalence of and factors associated with male perpetration of intimate partner violence: Findings from the UN multi-country cross-sectional study on men and violence in Asia and the Pacific. Lancet Glob Heal [Internet] Fulu et al. Open Access article distributed under the terms of CC BY-NC-ND; 2013;1(4):e187–e207. PMID:25104345

10. Postmus JL, Plummer SB, McMahon S, Murshid NS, Kim MS. Understanding Economic Abuse in the Lives of Survivors. J Interpers Violence 2012;27(3):411–430. PMID:21987509

11. Postmus JL, Hoge GL, Breckenridge J, Sharp-Jeffs N, Chung D. Economic Abuse as an Invisible Form of Domestic Violence: A Multicountry Review. Trauma, Violence, Abus 2020;21(2):261–283. PMID:29587598

12. Eriksson M, Ulmestig R. “It’s Not All About Money”: Toward a More Comprehensive Understanding of Financial Abuse in the Context of VAW. J Interpers Violence 2021;36(3–4):NP1625-1651NP. PMID:29295038

13. Karystianis G, Adily A, Schofield PW, Greenberg D, Jorm L, Nenadic G, Butler T. Automated analysis of domestic violence police reports to explore abuse types and victim injuries: Text mining study. J Med Internet Res 2019;21(3):1–12. PMID:30860490

14. Hoeboer C, de Roos C, van Son GE, Spinhoven P, Elzinga B. The effect of parental emotional abuse on the severity and treatment of PTSD symptoms in children and adolescents. Child Abus Negl [Internet] Elsevier Ltd; 2021;111(November 2020):104775. PMID:33158585

15. Hartanto A, Yong JC, Lee STH, Ng WQ, Tong EMW. Putting adversity in perspective: purpose in life moderates the link between childhood emotional abuse and neglect and adulthood depressive symptoms. J Ment Heal [Internet] Routledge; 2020;29(4):473–482. PMID:31983245

16. Karystianis G, Simpson A, Adily A, Schofield P, Greenberg D, Wand H, Nenadic G, Butler T. Prevalence of mental illnesses in domestic violence police records: Text mining study. J Med Internet Res 2020;22(12). PMID:33361056

17. Abramsky T, Devries K, Kiss L, Nakuti J, Kyegombe N, Starmann E, Cundill B, Francisco L, Kaye D, Musuya T, Michau L, Watts C. Findings from the SASA! Study: A cluster randomized controlled trial to assess the impact of a community mobilization intervention to prevent violence against women and reduce HIV risk in Kampala, Uganda. BMC Med 2014;12(1):15–17. PMID:25248996

18. Sylaska KM, Edwards KM. Disclosure of Intimate Partner Violence to Informal Social Support Network Members: A Review of the Literature. Trauma, Violence, Abus 2014;15(1):3–21. PMID:23887351

19. Thomas K, Modini T, Ringland V, Nancarrow H. Accurately identifying the ‘person most in need of protection’ in domestic and family violence law [Internet]. /Mnt/Conversions/Anrows/Files. 2020. Available from: https://anrows.intersearch.com.au/anrowsjspui/handle/1/18570ISBN:978-1-925925-62-3

20. Hamel JM. Perpetrator or victim? A review of the complexities of domestic violence cases. J Aggress Confl Peace Res 2020;12(2):55–62. [doi: 10.1108/JACPR-12-2019-0464]

21. Kourti A, Stavridou A, Panagouli E, Psaltopoulou T, Spiliopoulou C, Tsolia M, Sergentanis TN, Tsitsika A. Domestic Violence During the COVID-19 Pandemic: A Systematic Review. Trauma, Violence, Abus 2021; [doi: 10.1177/15248380211038690]

22. Walby S, Towers J, Balderston S, Corradi C, Francis B, Heiskanen M, Helweg-Larsen K, Mergaert L, Olive P, Palmer E, Stöckl H, Strid S. The concept and measurement of violence against women and men. concept Meas violence against women men. 2017. [doi: 10.26530/oapen_623150]ISBN:9781447332633

23. United Nations. Guidelines for Producing Statistics on Violence Against Women: Statistical Survey. United Nations Publ [Internet] 2014;1–208. Available from: https://unstats.un.org/unsd/gender/docs/Guidelines_Statistics_VAW.pdf

24. Jackson RG, Patel R, Jayatilleke N, Kolliakou A, Ball M, Gorrell G, Roberts A, Dobson RJ, Stewart R. Natural language processing to extract symptoms of severe mental illness from clinical text: The Clinical Record Interactive Search Comprehensive Data Extraction (CRIS-CODE) project. BMJ Open 2017;7(1):1–10. PMID:28096249

25. Liang J, Tsou C, Poddar A. A Novel System for Extractive Clinical Note Summarization using EHR Data. 2013;

26. Chandan JS, Thomas T, Bradbury-Jones C, Russell R, Bandyopadhyay S, Nirantharakumar K, Taylor J. Female survivors of intimate partner violence and risk of depression, anxiety and serious mental illness. Br J Psychiatry 2020;217(4):562–567. PMID:31171045

27. Estefan LF, Coulter ML, VandeWeerd C. Depression in Women Who Have Left Violent Relationships: The Unique Impact of Frequent Emotional Abuse. Violence Against Women 2016;22(11):1397–1413. PMID:26825117

28. Holliday, C. N., McCauley, H. L., Silverman, J. G., Ricci, E., Decker, M. R., Tancredi, D. J., & Miller, E. (2017). Racial/ethnic differences in women’s experiences of reproductive coercion, intimate partner violence, and unintended pregnancy. Journal of Women’s Health, 26(8), 828–835.

29. Ragavan, M. I., Thomas, K. A., Fulambarker, A., Zaricor, J., Goodman, L. A., & Bair-Merritt, M. H. (2020). Exploring the needs and lived experiences of racial and ethnic minority domestic violence survivors through community-based participatory research: A systematic review. Trauma, Violence, & Abuse, 21(5), 946–963.

30. Home Office. (2020). Draft statutory guidance: Framework for the new serious violence duty. UK Government. https://assets.publishing.service.gov.uk/government/uploads/system/uploads/attachment_data/file/896640/Draft_statutory_guidance_July_2020.pdf

31. UK Government. (2020). Domestic abuse draft statutory guidance framework [Chapter 2: Understanding domestic abuse]. GOV.UK. https://www.gov.uk/government/consultations/domestic-abuse-act-statutory-guidance/domestic-abuse-draft-statutory-guidance-framework#chapter-2--understanding-domestic-abuse

